# Meat intake and cancer risk: prospective analyses in UK Biobank

**DOI:** 10.1101/19003822

**Authors:** Anika Knuppel, Keren Papier, Georgina K. Fensom, Paul N. Appleby, Julie A. Schmidt, Tammy Y. N. Tong, Ruth C. Travis, Timothy J. Key, Aurora Perez-Cornago

**Affiliations:** Cancer Epidemiology Unit, Nuffield Department of Population Health, University of Oxford, Oxford, United Kingdom

## Abstract

**Background:** Red and processed meat has been consistently associated with risk for colorectal cancer, but evidence for other cancer sites is limited and few studies have examined the association between poultry intake and cancer risk. We examined associations between total meat, red meat, processed meat and poultry intake and incidence for 20 common cancer sites.

**Methods and Findings:** We analysed data from 475,023 participants (54% women) in UK Biobank. Participants were aged 37-73 years and cancer free at baseline. Information on meat consumption was based on a touchscreen questionnaire completed at baseline covering type and frequency of meat intake. Diet intake was re-measured a minimum of three times in a subsample (15%) using a web-based 24h dietary recall questionnaire. Multivariable-adjusted Cox proportional hazards models were used to determine the association between baseline meat intake and cancer incidence. Trends in risk across baseline meat intake categories were calculated by assigning a mean value to each category using estimates from the re-measured meat intakes. During a mean follow-up of 6.9 years, 28,955 participants were diagnosed with a malignant cancer. Total, red and processed meat intakes were each positively associated with risk of colorectal cancer (e.g. hazard ratio (HR) per 70 g/day higher intake of red and processed meat combined 1.31, 95%-confidence interval (CI) 1.14-1.52).

Red meat intake was positively associated with breast cancer (HR per 50 g/day higher intake 1.12, 1.01-1.24) and prostate cancer (1.15, 1.03-1.29). Poultry intake was positively associated with risk for cancers of the lymphatic and hematopoietic tissues (HR per 30g/day higher intake 1.16, 1.03-1.32). Only the associations with colorectal cancer were robust to Bonferroni correction for multiple comparisons. Study limitations include unrepresentativeness of the study sample for the UK population, low case numbers for less common cancers and the possibility of residual confounding.

**Conclusions:** Higher intakes of red and processed meat were associated with a higher risk of colorectal cancer. The observed positive associations of red meat consumption with breast and prostate cancer, and poultry intake with cancers of the lymphatic and hematopoietic tissues, require further investigation.

## Introduction

In 2014, neoplasms overtook cardiovascular diseases as the leading cause of death in the UK [1]. In 2018, an estimated 446,942 UK citizens were newly diagnosed with cancer; the most common cancer sites were breast, prostate, lung, colorectal cancer and malignant melanoma [2]. Cancer costs the UK £5 billion a year in cancer services and a total of £18.3 billion when including the costs for loss of productivity [3]. Therefore, identifying risk factors that may be targeted for cancer prevention is of great importance.

The latest meta-analysis from the World Cancer Research Fund (WCRF) / American Institute for Cancer Research (AICR) concluded that red meat was a probable cause and processed meat a convincing cause of colorectal cancer [4]. However, evidence for associations between red and processed meat intake and cancers at sites other than the colorectum was limited [4]. Furthermore, no conclusions on poultry intake and cancer risk were reached, mainly due to a low number of studies meeting the quality criteria [4]. Similarly, an International Agency for Research on Cancer (IARC) Monographs expert Working Group has classified red meat as Group 2A, *probably carcinogenic to humans* and processed meat as Group 1, *carcinogenic to humans* [5]. The IARC review also found some evidence of positive associations with consumption of red meat for cancers of the pancreas and prostate [5]. The interpretation of findings from published studies, however, is challenging due to the heterogeneity in the findings between individual studies, which may be the result of differences in exposure definitions, such as what constitutes processed meat, differences in dietary measurement, the handling of confounders and cohort specifics such as length of follow-up or sample size [6]. By investigating the association between meat intake and multiple cancer sites in the same cohort, definitions and analyses can be standardised and bias in outcome selection based on the results can be eliminated [7-10].

The aim of the present study was to systematically investigate the associations between intakes of total, red and processed meat (both separately and combined) and poultry and cancer across 20 common cancer sites in a large British cohort.

## Methods

### Study population

The UK Biobank study was established to investigate risk factors for major diseases in middle and older age. Between 2006 and 2010, 9.2 million individuals registered with the UK Health Service in England, Wales and Scotland were invited to participate in the study [11, 12]. In total, 503,317 men and women aged 37-73 years consented to participate and attended the baseline assessment [13]. The study was approved by The National Information Governance Board for Health and Social Care and the National Health Service North West Multicentre Research Ethics Committee (reference number 06/MRE08/65), and participants provided informed consent at baseline and to be followed up using data-linkage. By 19 March 2019 781 participants had withdrawn their consent to remain in the study.

### Exposure assessment

Data on dietary intake were collected at recruitment using 29 questions in a self-report touchscreen questionnaire (http://biobank.ctsu.ox.ac.uk/showcase/docs/TouchscreenQuestionsMainFinal.pdf). In the touchscreen questionnaire, meat intake was based on five questions concerning processed meat (including both red and white processed meat), and unprocessed meat (poultry, beef, lamb/mutton and pork). Answer options ranged from *‘never’, ‘less than once a week’, ‘once a week’, ‘2-4 times a week’, ‘5-6 times a week’* to *‘once or more daily’*, as well as *‘do not know’* and *‘prefer not to answer’*. The answer options were coded as intakes of 0, 0.5, 1, 3, 5.5 and 7 times/week. Intakes of unprocessed beef, lamb/mutton and pork were combined and classified as red meat intake. To investigate additive effects, red and processed meat intakes were combined as red and processed meat intake, and all meat types including poultry were combined as total meat intake. Meat intakes were categorised into three or four groups depending on distribution and restricted by the answer categories as follows: red meat 0 to <1, 1 to <2, 2 to <4 and ≥ 4 times/week; processed meat 0 to <1 time/week, 1 to <2 times a week and ≥ 2 times/week; poultry 0 to <1, 1 to <2 and ≥ 2 times/week; red and processed meat 0 to 1, >1 to <3, 3 to <5 and ≥ 5 times/week; total meat 3, 3 to <5, 5 to <7 and ≥7 times/week.

From 2009 onwards, a subsample of participants filled out an additional web-based 24h dietary recall questionnaire (the Oxford WebQ) [14, 15]. Mean meat intakes in each category were derived from data in a subsample of participants that had participated repeatedly (minimally 3 times, participant specific means were averaged over the repeated measurements) in the Oxford WebQ and were without a cancer diagnosis at time of data collection (n = 69,076) [14, 16, 17]. These average intakes in the categories for the subsample were then assigned to the intake categories in all participants to calculate trends in risk. Further details on how diet was collected and grouped can be found in S1 Text.

### Outcome assessment

Data on cancer diagnoses were provided by the Medical Research Information Service of the National Health Service (NHS) Information centre (for participants resident in England or Wales) and the Information Services Division of NHS Scotland (for participants resident in Scotland) [18]. The endpoints were first incident cancer diagnosis or cancer recorded in death certificates if no prior diagnosis was registered (all coded using the 10^th^ revision of the World Health Organization’s International Statistical Classification of Diseases (ICD-10) codes): oral (C00-14), oesophagus (C15), stomach (C16), colorectum (C18-20) including colon (C18) and rectum (including rectosigmoid junction; C19-20), liver (C22), pancreas (C25), lung (C34), malignant melanoma (C43), breast in women (C50), endometrium (C54), ovary (C56), prostate (C61), kidney (C64-65), bladder (C67), lymphatic and hematopoietic tissues (C81-96) and the subgroups non-Hodgkin lymphoma (C82-85), multiple myeloma (C90) and leukaemia (C91-95).

### Statistical analysis

Of 502,536 participants, 27,177 were excluded due to a cancer diagnosis at baseline (excluding non-melanoma skin cancer ICD-10: C44). Additionally, 334 participants were excluded in whom genetic sex differed from reported gender and two participants with zero person years of follow-up, resulting in a maximal study sample of 475,023 participants. Participants who reported ‘prefer not to answer’ or who did not know their meat intake were considered to have missing meat intake and were excluded from the respective analyses (for total meat 1.5%, red and processed meat 1.4%, red meat 1.3%, processed meat 0.4%, and poultry 0.3%). Missing data for the main covariates was minimal with 73.5% having no missing data and 94.5% having one or fewer covariates missing; therefore a ‘missing’ category was created for each covariate.

Each cancer site of interest was treated as a different endpoint for Cox proportional hazards regressions with age as the underlying time variable. The person-years of follow-up were calculated from baseline assessment until the first registration of malignant cancer, date of death due to cancer if not diagnosed previously, date of death, loss of follow-up or end of follow-up (31 March 2016 for England and Wales, 31 October 2015 for Scotland), whichever came first. Meat intakes were included in the regression models categorically using the lowest intake category as reference, and continuously as a trend in risk expressing the hazard ratios (HR) in increments of 100 g/day for total meat intake, 70 g/day for red and processed meat, 50 g/day for red meat, 20 g/day for processed meat and 30 g/day for poultry (for more details see S1 Text). Covariates were chosen based on the literature for each cancer site, and availability of data at baseline.

Minimally adjusted models (Model 0) were stratified by geographical region of baseline assessment centre (6 regions: London, Wales, North-West, North-East, Yorkshire and Humber, West Midlands, East Midlands, South-East, South-West, Scotland), sex, and age group at recruitment (<45, 45-<50, 50-<55, 55-<60, 60-<65 and ≥65 years). Subsequently, multivariable models were adjusted for ethnicity (four groups where possible: White, Asian or Asian British, Black or Black British, and Mixed Race or Other; and two groups: White, Non-White for oral, oesophagus, stomach cancer and malignant melanoma due to low numbers of cases for Non-White ethnicity; unknown), Townsend deprivation score (quintiles, unknown), educational qualifications (Professional qualifications/NVQ/HND/HNC/Degree or other professional qualification referred to as College or university degree/vocational qualification; A levels or Scottish Highers referred to as national examination at ages 17-18; O level/GCSEs/CSEs referred to as national examination at age 16; other/unknown), employment status (in paid employment, pension, not in paid employment, unknown), living with a spouse or partner (yes, no, unknown), height (sex-specific quintiles, unknown), smoking (never, former, current <15 cigarettes/day, current ≥ 15 cigarettes/day, current amount unknown, unknown), physical activity (low, moderate, high, unknown), alcohol intake (non-drinker, <1 g/day, 1 to <10 g/day, 10 to <20 g/day, ≥20 g/day, unknown), total fruit and vegetable intake (<3 servings/day, 3 to <4 servings/day, 4 to <6 servings/day, ≥ 6 servings/day, unknown), and estimated cereal fibre intake (sex-specific quintiles, unknown) (Model 1) [16]. In women, Model 1 was additionally adjusted for menopausal status (pre-, postmenopausal, unknown), parity (nulliparous, 1 to 2, ≥ 3, unknown), hormone-replacement therapy (HRT) (never, past, current, unknown) and oral contraceptive pill use (OCP) (never, past, current, unknown). Model 2 was additionally adjusted for measured body mass index (BMI) (sex-specific quintiles, unknown). Analyses for meat intake and melanoma skin cancer were additionally adjusted for skin colour (very fair, fair, light olive, dark olive, brown/black, unknown), hair colour (blonde, red, light brown, dark brown, black, unknown), skin reaction to sun exposure (get very tanned, moderately tanning, mildly/occasionally tanning, never tanning only burning, unknown), sun or UV protection use (never/rarely, sometimes, most of the time, always, do not go out in the sunshine, unknown), sunburns before age 15 (never, ever, unknown) and solarium use (never, 1 to 2 times/year, ≥3 times/year, unknown). Detailed information on data-collection and categorisation of covariates is given in the S2 Text.

We conducted three sensitivity analyses: (1) we repeated all analyses excluding the first 2 years of follow-up, to examine whether the overall results might be influenced by reverse causality; (2) we repeated analyses including only those reporting never having smoked at baseline, to reduce residual confounding by smoking; (3) we compared models with and without an interaction term by sex using likelihood-ratio tests, based on earlier findings of sex differences in the associations between meat intake and cancer risk in this cohort [19].

Statistical significance was set at the 5% level and for hazard ratios (HRs) 95% confidence intervals (CIs) were calculated. All analyses were performed using Stata release 15.1 [20].

## Results

### Descriptive

During a mean follow-up of 6.9 (SD 1.3, maximum 10.1) years, a total of 28,955 participants (6.1%) were newly diagnosed with any type of malignant cancer (excluding non-melanoma skin cancer ICD-10: C44). Table 1 shows characteristics of all participants and those who developed a cancer during follow-up. Participants who developed cancer were older, less physically active and had a higher BMI compared to participants as a whole, more likely to be retired, current or former smokers and heavy alcohol drinkers. Women who developed cancer were more likely to be postmenopausal and to have used HRT. Participants reporting higher meat intakes were more likely to be men, from areas with higher affluence (measured by Townsend score), living with a spouse or partner, former or current smokers, less physically active, to drink more alcohol, to report lower fruit, vegetable and cereal fibre intake, and had a higher BMI (see S2-3 Tables for baseline characteristics by intakes of red and processed meat and poultry).

**Table 1.** Baseline characteristics of included UK Biobank participants.

| Characteristics* | All participants (n = 475,023) | Participants who developed any malignant cancer (n = 28,955) |
| --- | --- | --- |
| **Sociodemographic** |  |  |
| Sex, n (%) |  |  |
| Women | 255,974 (53.9) | 13,832 (47.8) |
| Men | 219,049 (46.1) | 15,123 (52.2) |
| Age (years) | 56.3 (8.1) | 60.2 (6.8) |
| Ethnicity, n (%) |  |  |
| White | 446,191 (93.9) | 27,788 (96.0) |
| Asian or Asian British | 11,156 (2.3) | 394 (1.4) |
| Black or Black British | 7,776 (1.6) | 303 (1.0) |
| Mixed Race or other | 7,243 (1.5) | 303 (1.0) |
| Unknown | 2,657 (0.6) | 167 (0.6) |
| Townsend deprivation, n (%) |  |  |
| Most affluent (mean -4.7) | 95,023 (20.0) | 5,875 (20.3) |
| 2 (mean -3.3) | 94,823 (20.0) | 5,920 (20.4) |
| 3 (mean -2.1) | 94,817 (20.0) | 5,782 (20.0) |
| 4 (mean -0.9) | 94,885 (20.0) | 5,625 (19.4) |
| Most deprived (mean 3.8) | 94,881 (20.0) | 5,729 (19.8) |
| Unknown | 594 (0.1) | 24 (0.1) |
| Qualification, n (%) |  |  |
| College or university degree/vocational qualification | 281,507 (59.3) | 15,840 (54.7) |
| National examination at ages 17-18 | 25,815 (5.4) | 1,430 (4.9) |
| National examination at age 16 | 78,724 (16.6) | 4,448 (15.4) |
| Other/unknown | 88,977 (18.7) | 7,237 (25.0) |
| Employment, n (%) |  |  |
| In paid employment | 276,136 (58.1) | 12,902 (44.6) |
| Pension | 140,140 (29.5) | 12,535 (43.3) |
| Not in paid employment | 53,273 (11.2) | 3,236 (11.2) |
| Unknown | 5,474 (1.2) | 282 (1.0) |
| Living with a spouse/partner, n (%) |  |  |
| Living with partner, | 343,752 (72.4) | 20,955 (72.4) |
| Not living with partner | 129,249 (27.2) | 7,891 (27.3) |
| Unknown | 2,022 (0.4) | 109 (0.4) |
| **Anthropometric** |  |  |
| Standing height (cm) in women | 162.4 (6.3) | 162.5 (6.3) |
| Standing height (cm) in men | 175.6 (6.8) | 175.2 (6.8) |
| Body mass index (kg/m<sup>2</sup>) | 27.4 (4.8) | 27.8 (4.8) |
| **Lifestyle** |  |  |
| Smoking, n (%) |  |  |
| Never | 259,528 (54.6) | 13,508 (46.7) |
| Former | 162,277 (34.2) | 11,482 (39.7) |
| Current <15 cigarettes/day | 14,475 (3.0) | 974 (3.4) |
| Current ≥15 cigarettes/day | 19,942 (4.2) | 1,737 (6.0) |
| Current, amount unknown | 16,035 (3.4) | 1,061 (3.7) |
| Unknown | 2,766 (0.6) | 193 (0.7) |
| Physical activity level, n (%) |  |  |
| High ≥50 excess METs | 75,481 (15.9) | 4,419 (15.3) |
| Moderate 10-<50 excess METs | 230,667 (48.6) | 13,731 (47.4) |
| Low <10 excess METs | 149,662 (31.5) | 9,576 (33.1) |
| Unknown | 19,213 (4.0) | 1,229 (4.2) |
| Alcohol intake, n (%) |  |  |
| None drinkers | 38,113 (8.0) | 2,309 (8.0) |
| <1 g/day | 53,265 (11.2) | 3,260 (11.3) |
| 1-<10 g/day | 148,406 (31.2) | 8,394 (29.0) |
| 10-<20 g/day | 102,297 (21.5) | 6,052 (20.9) |
| ≥20 g/day | 129,466 (27.3) | 8,733 (30.2) |
| Unknown | 3,476 (0.7) | 207 (0.7) |
| **Diet** |  |  |
| Fruit and vegetable intake (servings/day) | 4.69 (2.61) | 4.64 (2.49) |
| Estimated cereal fibre intake (g/day) | 4.50 (2.94) | 4.54 (2.93) |
| **Women's health** |  |  |
| Menopausal status, n (%) |  |  |
| Premenopausal | 60,795 (23.8) | 2,005 (14.5) |
| Postmenopausal | 180,492 (70.5) | 11,259 (81.4) |
| Unknown | 14,687 (5.7) | 568 (4.1) |
| Parity, n (%) |  |  |
| 0 births | 47,650 (18.6) | 2,565 (18.5) |
| 1-2 births | 145,721 (56.9) | 7,759 (56.1) |
| ≥3 births | 61,842 (24.2) | 3,470 (25.1) |
| Unknown | 761 (0.3) | 38 (0.3) |
| Hormone replacement therapy use, n (%) |  |  |
| Never | 158,225 (61.8) | 7,446 (53.8) |
| Past | 80,650 (31.5) | 5,291 (38.3) |
| Current | 15,612 (6.1) | 1,017 (7.4) |
| Unknown | 1,487 (0.6) | 78 (0.6) |
| Oral contraceptive pill use, n (%) |  |  |
| Never | 47,533 (18.6) | 3,035 (21.9) |
| Past | 202,257 (79.0) | 10,532 (76.1) |
| Current | 4,855 (1.9) | 203 (1.5) |
| Unknown | 1,329 (0.5) | 62 (0.4) |

BMI, Body mass index; MET, metabolic equivalents.
\* Mean (Standard deviation) unless otherwise specified.

Intakes of different meat types were positively correlated, with a high proportion of those reporting high intakes of one type of meat also reporting high intakes of other types. Of participants who reported high red meat intakes (≥ 4 times/week) 44.4% also reported high processed meat intakes (≥ 2 times/week) and only 2.6% no processed meat intake. Of participants reporting high intakes of poultry (3-<5 times/week) 30.6% also reported high intakes of red and processed meat (≥ 5 times/week), while only 4.6% reported eating red and processed meat ≤1 time/week (S3 Table).

### Main findings

Figs 1-5 depict estimated HRs of each individual cancer site associated with an incremental increase in meat intake for the fully adjusted models (Model 2). Tables S4-S8 show estimated HRs by meat intake categories and per incremental increase in intake at three or four levels of adjustment. We report trends in risk where associations showed similar directions to associations by intake categories, and only refer to categorical results if they differed.

**Fig 1.**
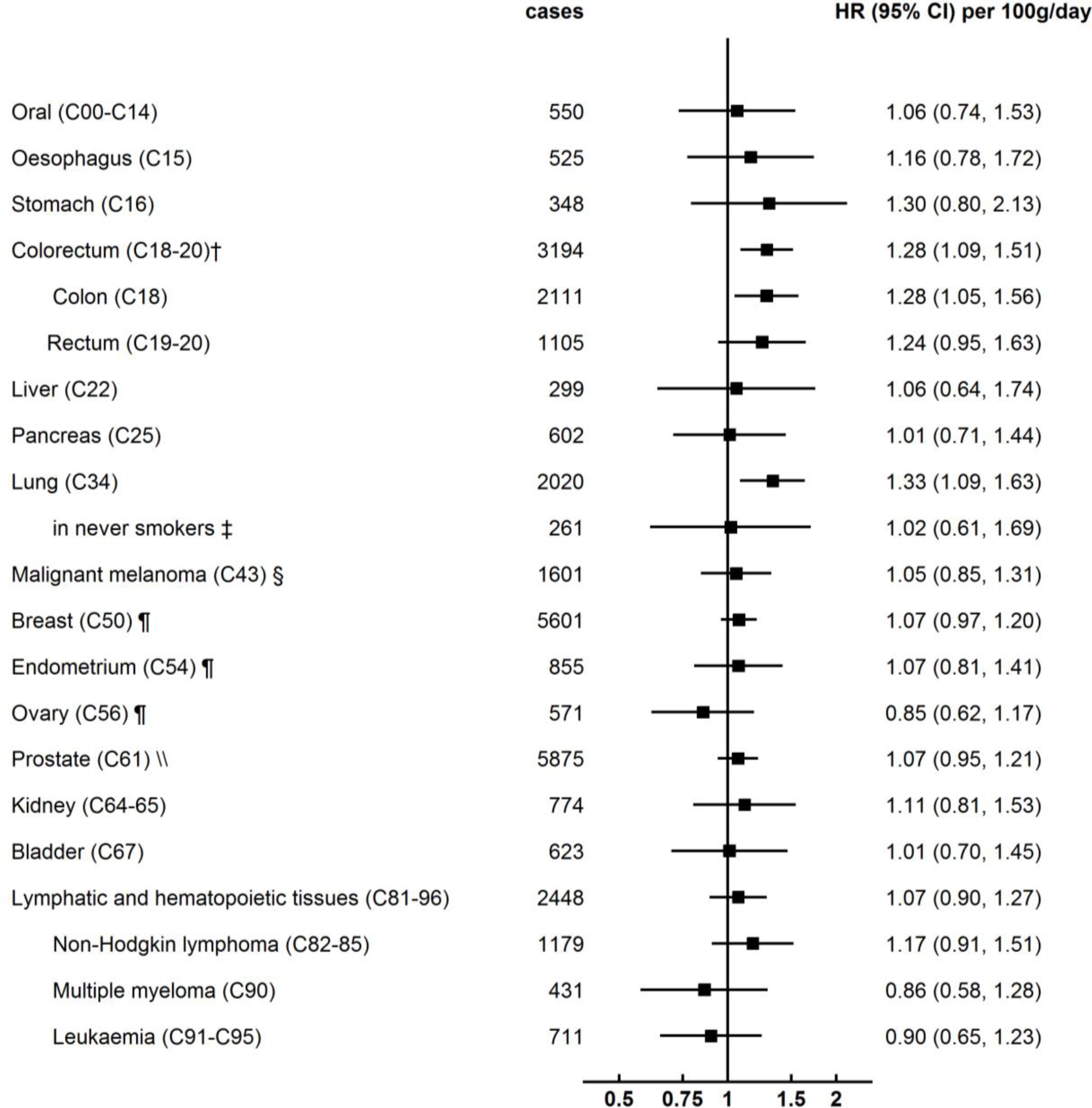
Association of total meat intake and cancer incidence by cancer site (per 100 g/day higher intake; n = 467,420)* CI, Confidence interval; HR, Hazard ratio. **\***Stratified for sex, age group (<45, 45-<50, 50-<55, 55-<60, 60-<65 and ≥65 years) and region (London, Wales, North-West, North-East, Yorkshire and Humber, West Midlands, East Midlands, South-East, South-West, Scotland) and adjusted for age (underlying time variable), ethnicity (4 groups where possible: White, Asian or Asian British, Black or Black British, Mixed race or other, unknown), deprivation (Townsend index quintiles, unknown), qualification (college or university degree/vocational qualification, national examination at ages 17-18, national examination at age 16, other/unknown), employment (in paid employment, receiving pension, not in paid employment, unknown), living with a spouse or partner (yes, no, unknown), height (sex-specific quintiles, unknown), smoking (never, former, current <15 cigarettes/day, current ≥15 cigarettes/day, current unknown amount of cigarettes/day, unknown), physical activity (<10 excess metabolic equivalents, 10-<50 excess metabolic equivalents, ≥50 excess metabolic equivalents, unknown), alcohol intake (none, <1 g/day, 1-<10 g/day, 10-<20 g/day, ≥20 g/day, unknown), total fruit and vegetable intake (<3 servings/day, 3-<4 servings/day, 4-<6 servings/day, ≥6 servings/day, unknown), estimated cereal fibre intake (sex-specific quintiles, unknown), body mass index (sex-specific quintiles, unknown), in women: menopausal status (pre-, postmenopausal, unknown), parity (nulliparous, 1-2, ≥3, unknown), hormone-replacement therapy (never, past, current, unknown) and oral contraceptive pill use (never, past, current, unknown). † In 22 cases of colorectal cancer (C18/20) colon (C18) and rectal cancer (C19-20) diagnoses coincided. ‡ Analyses restricted to never smokers (n = 256,009). § Additionally adjusted for skin colour (very fair, fair, light olive, dark olive, brown/black, unknown), hair colour (blonde, red, light brown, dark brown, black, other/unknown), skin reaction (get very tanned, moderately tanning, mildly/occasionally tanning, never tanning only burning, unknown), UV protection use (never/rarely, sometimes, most of the time, always, do not go out in the sunshine, unknown), sunburns before age 15 (never, ever, unknown), solarium use (never, >0-2 times/year, ≥3 times/year, unknown). ¶ Analyses restricted to women (n = 252,268). \\ Analyses restricted to men (n = 215,152).

Total meat intake was positively associated with the risk of colorectal cancer (HR per 100 g/day higher intake 1.28, 95%-CI 1.09-1.51, P_trend_ = 0.0022; Fig 1) and colon cancer (1.28, 1.05-1.56, P_trend_ = 0.013; Fig 1). There was a positive association between total meat intake and lung cancer risk, but not when current and former smokers were excluded from the analysis (Fig 1).

Red and processed meat intake was associated with a higher risk of colorectal cancer (HR per 70 g/day higher intake 1.33, 95%-CI 1.14-1.52, P_trend_ = 0.0002; Fig 2) and colon cancer (1.38, 1.16-1.66, P_trend_= 0.0004; Fig 2). There was a positive association between red and processed meat intake and lung cancer risk, but not after exclusion of former and current smokers (Fig 2). In analyses using red and processed meat intake categorically, there were additional associations with malignant melanoma and prostate cancer. Participants who consumed red and processed meat >1 to <3 or 3 to <5 times/week had a higher risk of malignant melanoma compared to those who consumed red and processed meat one or less times/week (HR for intakes >1-<3 times/week 1.27, 95%-CI 1.02-1.58; for intakes 3-<5 times/week 1.40, 1.12-1.74; S5 Table); the risk did not differ in participants who consumed red and processed meat ≥ 5 times/week (HR 1.17, 95%-CI 0.93-1.47; S5 Table). Any red and processed meat intake above one time/week was associated with a higher risk of prostate cancer (HR for intakes >1-<3 times/week 1.18, 95%-CI 1.03-1.34; for intakes 3-<5 times/week 1.18, 1.03-1.35; for intakes ≥ 5 times/week 1.15, 1.01-1.31; reference intake ≤1 time/week; S5 Table).

**Fig 2.**
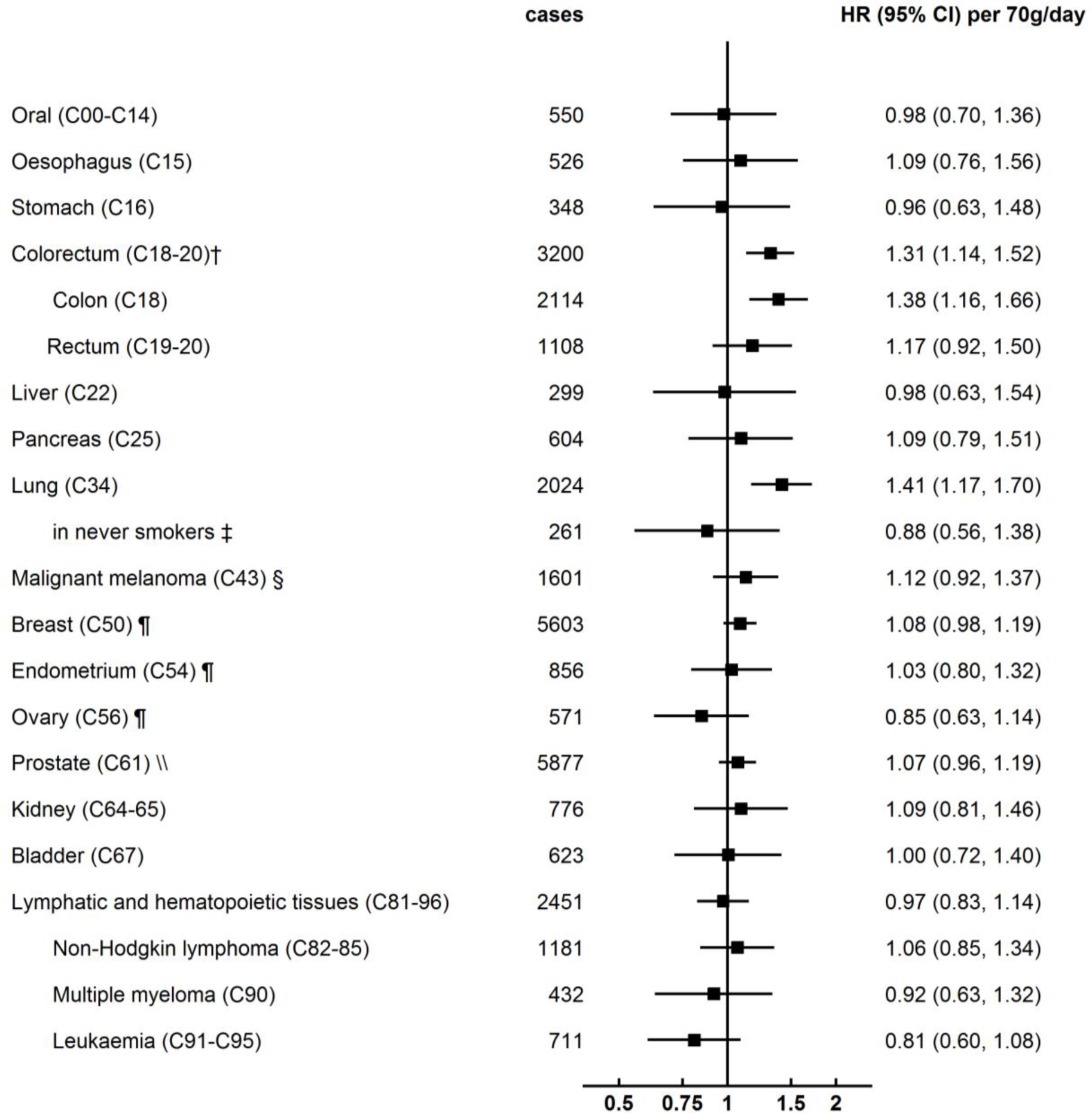
Association of red and processed meat intake and cancer incidence by cancer site (per 70 g/day higher intake; n = 467,777)* CI, Confidence interval; HR, Hazard ratio. **\***Stratified for sex, age group (<45, 45-<50, 50-<55, 55-<60, 60-<65 and ≥65 years) and region (London, Wales, North-West, North-East, Yorkshire and Humber, West Midlands, East Midlands, South-East, South-West, Scotland) and adjusted for age (underlying time variable), ethnicity (4 groups where possible: White, Asian or Asian British, Black or Black British, Mixed race or other, unknown), deprivation (Townsend index quintiles, unknown), qualification (college or university degree/vocational qualification, national examination at ages 17-18, national examination at age 16, other/unknown), employment (in paid employment, receiving pension, not in paid employment, unknown), living with a spouse or partner (yes, no, unknown), height (sex-specific quintiles, unknown), smoking (never, former, current <15 cigarettes/day, current ≥15 cigarettes/day, current unknown amount of cigarettes/day, unknown), physical activity (<10 excess metabolic equivalents, 10-<50 excess metabolic equivalents, ≥50 excess metabolic equivalents, unknown), alcohol intake (none, <1 g/day, 1-<10 g/day, 10-<20 g/day, ≥20 g/day, unknown), total fruit and vegetable intake (<3 servings/day, 3-<4 servings/day, 4-<6 servings/day, ≥6 servings/day, unknown), estimated cereal fibre intake (sex-specific quintiles, unknown), body mass index (sex-specific quintiles, unknown), in women: menopausal status (pre-, postmenopausal, unknown), parity (nulliparous, 1-2, ≥3, unknown), hormone-replacement therapy (never, past, current, unknown) and oral contraceptive pill use (never, past, current, unknown). † In 22 cases of colorectal cancer (C18/20) colon (C18) and rectal cancer (C19-20) diagnoses coincided. ‡ Analyses restricted to never smokers (n = 256,182). § Additionally adjusted for skin colour (very fair, fair, light olive, dark olive, brown/black, unknown), hair colour (blonde, red, light brown, dark brown, black, other/unknown), skin reaction (get very tanned, moderately tanning, mildly/occasionally tanning, never tanning only burning, unknown), UV protection use (never/rarely, sometimes, most of the time, always, do not go out in the sunshine, unknown), sunburns before age 15 (never, ever, unknown), solarium use (never, >0-2 times/year, ≥3 times/year, unknown). ¶ Analyses restricted to women (n = 252,422). \\ Analyses restricted to men (n = 215,355).

Red meat intake was associated with a higher risk of colorectal cancer (HR per 50 g/day higher intake 1.22, 95%-CI 1.05-1.42, P_trend_ = 0.0009; Fig 3), colon cancer (1.34, 1.11-1.61, P_trend_ = 0.0021; Fig 3), breast cancer in women (1.12, 1.02-1.24, P_trend_ = 0.030; Fig 3) and prostate cancer in men (1.16, 1.03-1.30, P_trend_ = 0.013; Fig 3). Additional adjustment for enlarged prostate at baseline did not materially change the association between red meat intake and prostate cancer risk (S6 Table). There was a positive association between red meat intake and lung cancer, but not after the exclusion of participants who were current or former smokers (Fig 3). In analyses using red meat intake categorically, participants who consumed 1 to <2 or 2 to <3 times/week had higher risks of malignant melanoma compared to participants who consumed red meat less than one time per week (HR for intakes 1-<2 times/week 1.32, 95%-CI 1.07-1.62; for intakes 2-<3 times/week 1.31, 1.07-1.62; S6 Table); the risk did not differ in participants who consumed red meat ≥ 3 times/week (1.09, 95%-CI 0.85-1.39; S6 Table).

**Fig 3.**
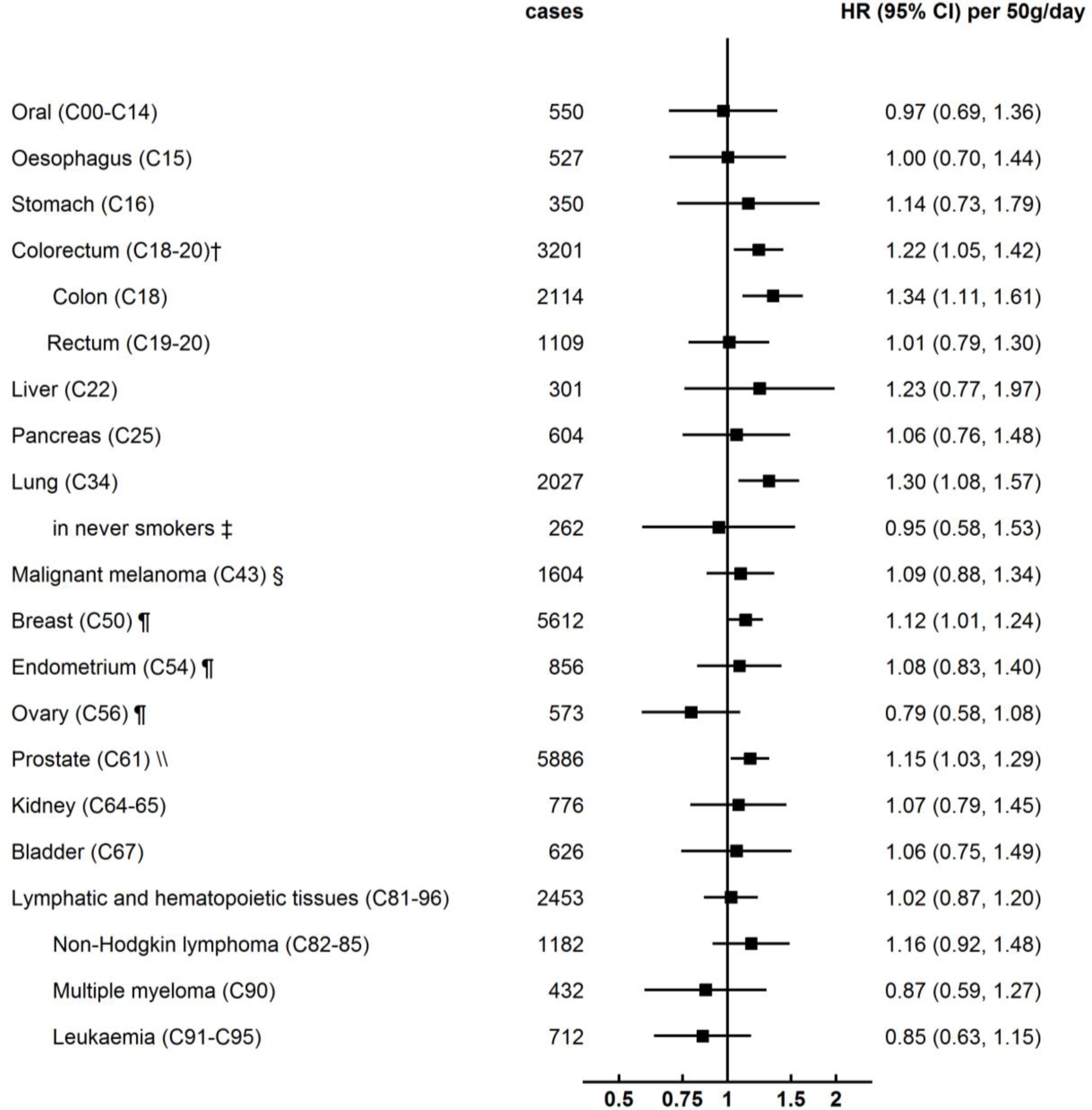
Association of red meat intake and cancer incidence by cancer site (per 50 g/day higher intake; n = 468,364)* CI, Confidence interval; HR, Hazard ratio. **\***Stratified for sex, age group (<45, 45-<50, 50-<55, 55-<60, 60-<65 and ≥65 years) and region (London, Wales, North-West, North-East, Yorkshire and Humber, West Midlands, East Midlands, South-East, South-West, Scotland) and adjusted for age (underlying time variable), ethnicity (4 groups where possible: White, Asian or Asian British, Black or Black British, Mixed race or other, unknown), deprivation (Townsend index quintiles, unknown), qualification (college or university degree/vocational qualification, national examination at ages 17-18, national examination at age 16, other/unknown), employment (in paid employment, receiving pension, not in paid employment, unknown), living with a spouse or partner (yes, no, unknown), height (sex-specific quintiles, unknown), smoking (never, former, current <15 cigarettes/day, current ≥15 cigarettes/day, current unknown amount of cigarettes/day, unknown), physical activity (<10 excess metabolic equivalents, 10-<50 excess metabolic equivalents, ≥50 excess metabolic equivalents, unknown), alcohol intake (none, <1 g/day, 1-<10 g/day, 10-<20 g/day, ≥20 g/day, unknown), total fruit and vegetable intake (<3 servings/day, 3-<4 servings/day, 4-<6 servings/day, ≥6 servings/day, unknown), estimated cereal fibre intake (sex-specific quintiles, unknown), body mass index (sex-specific quintiles, unknown), in women: menopausal status (pre-, postmenopausal, unknown), parity (nulliparous, 1-2, ≥3, unknown), hormone-replacement therapy (never, past, current, unknown) and oral contraceptive pill use (never, past, current, unknown). † In 22 cases of colorectal cancer (C18/20) colon (C18) and rectal cancer (C19-20) diagnoses coincided. ‡ Analyses restricted to never smokers (n = 256,545). § Additionally adjusted for skin colour (very fair, fair, light olive, dark olive, brown/black, unknown), hair colour (blonde, red, light brown, dark brown, black, other/unknown), skin reaction (get very tanned, moderately tanning, mildly/occasionally tanning, never tanning only burning, unknown), UV protection use (never/rarely, sometimes, most of the time, always, do not go out in the sunshine, unknown), sunburns before age 15 (never, ever, unknown), solarium use (never, >0-2 times/year, ≥3 times/year, unknown). ¶ Analyses restricted to women (n = 252,745). \\ Analyses restricted to men (n = 215,619).

Processed meat intake was positively associated with the risk of colorectal cancer (HR per 20 g/day higher intake 1.18, 95%-CI 1.06-1.31, P_trend_ = 0.0022; Fig 4) and rectal cancer (1.26, 1.06-1.51, P_trend_ = 0.011; Fig 4). Processed meat intake was positively associated with lung cancer, but not in participants who had never smoked (Fig 4). In analyses using processed meat intake categorically, participants who consumed processed meat 1 to <2 times/week but not higher, had higher risks of breast cancer compared to participants who consumed no processed meat (HR 1.11, 95%-CI 1.01-1.21; for intakes >0-<1 time/week 1.06, 0.97-1.16; for intakes ≥ 2 times/week 1.07, 0.97-1.19; S7 Table).

**Fig 4.**
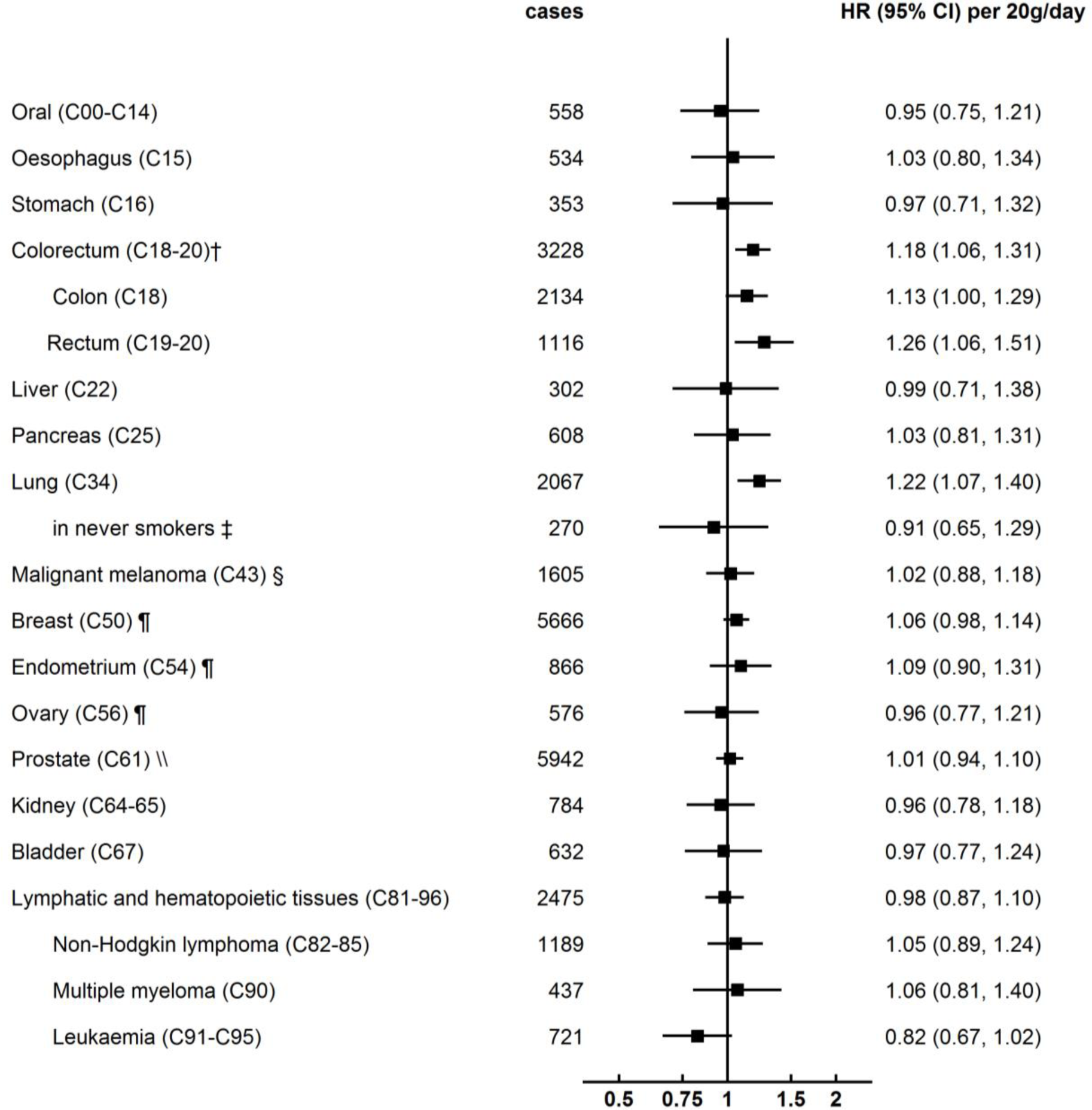
Association of processed meat intake and cancer incidence by cancer site (per 20 g/day higher intake; n = 472,881)* CI, Confidence interval; HR, Hazard ratio. **\***Stratified for sex, age group (<45, 45-<50, 50-<55, 55-<60, 60-<65 and ≥65 years) and region (London, Wales, North-West, North-East, Yorkshire and Humber, West Midlands, East Midlands, South-East, South-West, Scotland) and adjusted for age (underlying time variable), ethnicity (4 groups where possible: White, Asian or Asian British, Black or Black British, Mixed race or other, unknown), deprivation (Townsend index quintiles, unknown), qualification (college or university degree/vocational qualification, national examination at ages 17-18, national examination at age 16, other/unknown), employment (in paid employment, receiving pension, not in paid employment, unknown), living with a spouse or partner (yes, no, unknown), height (sex-specific quintiles, unknown), smoking (never, former, current <15 cigarettes/day, current ≥15 cigarettes/day, current unknown amount of cigarettes/day, unknown), physical activity (<10 excess metabolic equivalents, 10-<50 excess metabolic equivalents, ≥50 excess metabolic equivalents, unknown), alcohol intake (none, <1 g/day, 1-<10 g/day, 10-<20 g/day, ≥20 g/day, unknown), total fruit and vegetable intake (<3 servings/day, 3-<4 servings/day, 4-<6 servings/day, ≥6 servings/day, unknown), estimated cereal fibre intake (sex-specific quintiles, unknown), body mass index (sex-specific quintiles, unknown), in women: menopausal status (pre-, postmenopausal, unknown), parity (nulliparous, 1-2, ≥3, unknown), hormone-replacement therapy (never, past, current, unknown) and oral contraceptive pill use (never, past, current, unknown). † In 22 cases of colorectal cancer (C18/20) colon (C18) and rectal cancer (C19-20) diagnoses coincided. ‡ Analyses restricted to never smokers (n = 258,827). § Additionally adjusted for skin colour (very fair, fair, light olive, dark olive, brown/black, unknown), hair colour (blonde, red, light brown, dark brown, black, other/unknown), skin reaction (get very tanned, moderately tanning, mildly/occasionally tanning, never tanning only burning, unknown), UV protection use (never/rarely, sometimes, most of the time, always, do not go out in the sunshine, unknown), sunburns before age 15 (never, ever, unknown), solarium use (never, >0-2 times/year, ≥3 times/year, unknown). ¶ Analyses restricted to women (n = 254,909). \\ Analyses restricted to men (n = 217,972).

Poultry intake was associated with a higher risk of cancers of lymphatic and hematopoietic tissues (HR per 30 g/day higher intake 1.16, 95%-CI 1.03-1.32, P_trend_ = 0.019; Fig 5). In categorical analyses, participants who consumed poultry once or more times/week had a higher risk of prostate cancer (HR for intakes 1 to <2 times/week 1.11, 95%-CI 1.03-1.20; for intakes ≥ 2 times/week 1.09, 1.00-1.17; S8 Table) compared to participants who consumed poultry less than once a week.

**Fig 5.**
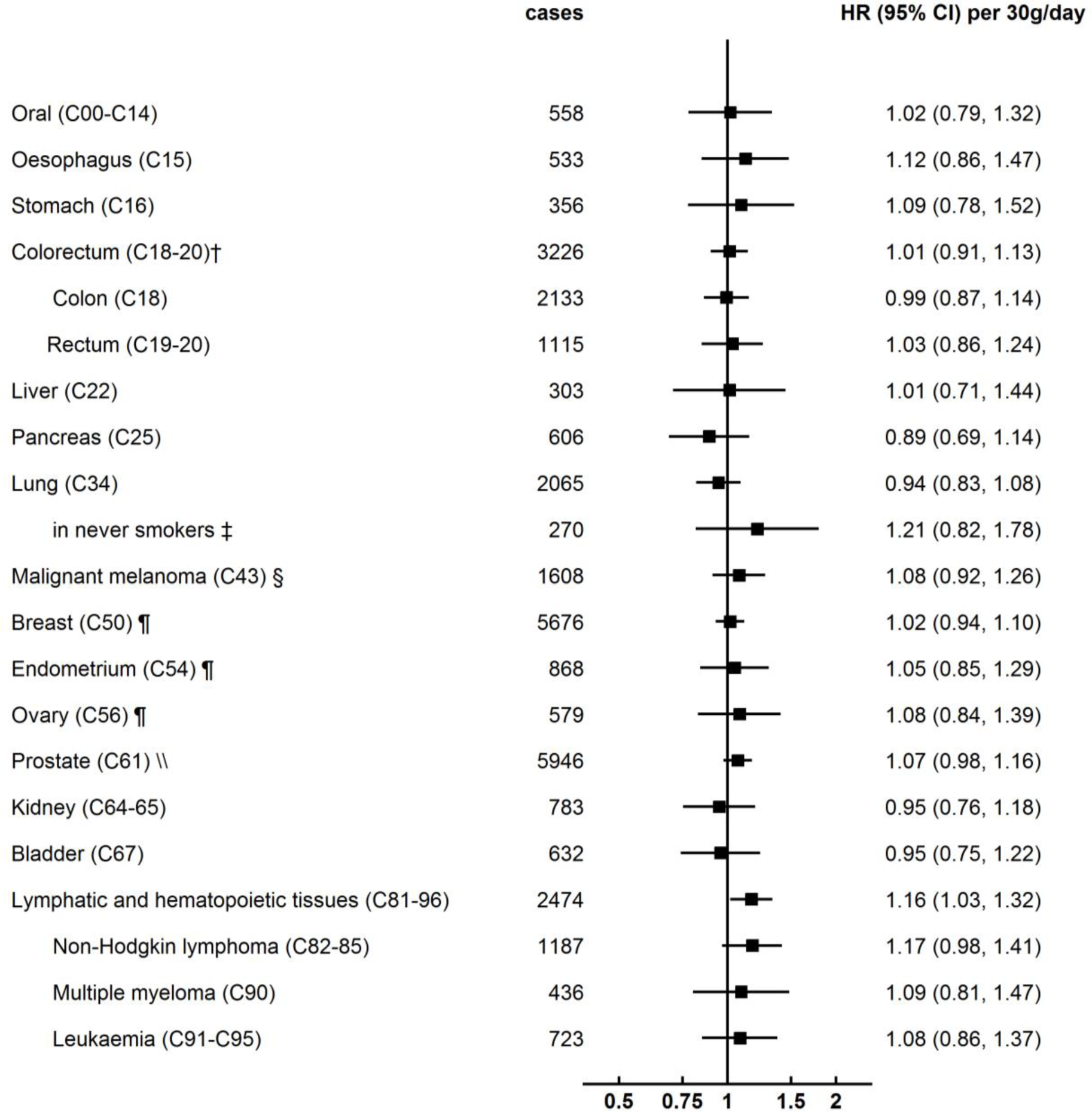
Association of poultry intake and cancer incidence by cancer site (per 30 g/day higher intake; n = 473,048)* CI, Confidence interval; HR, Hazard ratio. **\***Stratified for sex, age group (<45, 45-<50, 50-<55, 55-<60, 60-<65 and ≥65 years) and region (London, Wales, North-West, North-East, Yorkshire and Humber, West Midlands, East Midlands, South-East, South-West, Scotland) and adjusted for age (underlying time variable), ethnicity (4 groups where possible: White, Asian or Asian British, Black or Black British, Mixed race or other, unknown), deprivation (Townsend index quintiles, unknown), qualification (college or university degree/vocational qualification, national examination at ages 17-18, national examination at age 16, other/unknown), employment (in paid employment, receiving pension, not in paid employment, unknown), living with a spouse or partner (yes, no, unknown), height (sex-specific quintiles, unknown), smoking (never, former, current <15 cigarettes/day, current ≥15 cigarettes/day, current unknown amount of cigarettes/day, unknown), physical activity (<10 excess metabolic equivalents, 10-<50 excess metabolic equivalents, ≥50 excess metabolic equivalents, unknown), alcohol intake (none, <1 g/day, 1-<10 g/day, 10-<20 g/day, ≥20 g/day, unknown), total fruit and vegetable intake (<3 servings/day, 3-<4 servings/day, 4-<6 servings/day, ≥6 servings/day, unknown), estimated cereal fibre intake (sex-specific quintiles, unknown), body mass index (sex-specific quintiles, unknown), in women: menopausal status (pre-, postmenopausal, unknown), parity (nulliparous, 1-2, ≥3, unknown), hormone-replacement therapy (never, past, current, unknown) and oral contraceptive pill use (never, past, current, unknown). † In 22 cases of colorectal cancer (C18/20) colon (C18) and rectal cancer (C19-20) diagnoses coincided. ‡ Analyses restricted to never smokers (n = 259,040). § Additionally adjusted for skin colour (very fair, fair, light olive, dark olive, brown/black, unknown), hair colour (blonde, red, light brown, dark brown, black, other/unknown), skin reaction (get very tanned, moderately tanning, mildly/occasionally tanning, never tanning only burning, unknown), UV protection use (never/rarely, sometimes, most of the time, always, do not go out in the sunshine, unknown), sunburns before age 15 (never, ever, unknown), solarium use (never, >0-2 times/year, ≥3 times/year, unknown). ¶ Analyses restricted to women (n = 255,064). \\ Analyses restricted to men (n = 217,984).

### Sensitivity analyses

Associations were largely unchanged when excluding the first 2 years of follow-up (S1 - S5 Figs). Nonetheless, there was an additional association between total meat intake and a higher risk for stomach cancer (HR per 100 g/day higher total meat intake 1.82, 95%-CI 1.00-3.29; S1 Fig). In analyses restricted to never smokers most results were unchanged except for the analyses of lung cancer (S1 - S5 Figs). Still, in never smokers total and red meat intake were additionally positively associated with rectal cancer risk (HR per 100 g/day higher intake of total meat 1.84, 95%-CI 1.20-2.81, P_trend_ = 0.0051; S1 Fig; per 50 g/day higher intake of red meat 1.53, 1.04-2.25, P_trend_ = 0.032; S3 Fig). Furthermore, red meat intake was additionally inversely associated with ovarian cancer risk (HR per 50 g/day higher intake 0.63, 95%-CI 0.43-0.94, P_trend_ = 0.022; S3 Fig), processed meat intake was no longer associated with the risk of colon cancer (per 20 g/day higher intake of processed meat 1.01, 0.84-1.21, P_trend_ = 0.91; S4 Fig), and poultry was no longer associated with risk of cancers of lymphatic and hematopoietic tissues (per 30 g/day higher poultry intake 1.14, 0.95-1.36, P_trend_ = 0.17; S5 Fig).

There was no evidence for heterogeneity by sex except for associations with colorectal, colon and rectal cancer. While in men red and processed meat intake was associated with a higher risk of colorectal cancer (HR per 70 g higher red and processed meat intake 1.51, 95%-CI 1.22-1.86, P_trend_ = 0.0001), there was no association in women (1.16, 0.95-1.42, P_trend_ = 0.15; P_heterogeneity_ = 0.0073; S9 Table). There was similarly heterogeneity by sex in the association of red and processed meat with rectal cancer, with no association in women (in women: 0.91, 0.63-1.32, P_trend_ = 0.62; in men: 1.44, 1.01, 1.95, P_trend_ = 0.042; P_heterogeneity_ = 0.029; S9 Table). The heterogeneity by sex was less marked for associations with colon cancer (in women: 1.26, 0.99-1.60, P_trend_ = 0.057; in men: 1.59, 1.21, 2.09, P_trend_ = 0.0009; P_heterogeneity_ = 0.040, S9 Table). Additionally there was a weak inverse association between poultry intake and rectal cancer in women and a positive association in men (HR per 30 g higher poultry intake in women: 0.73, 95%-CI 0.54-0.99, P_trend_ = 0.046; in men: 1.27, 1.00-1.61, P_trend_ = 0.049; P_heterogeneity_ = 0.0033, S9 Table).

## Discussion

In this large British prospective study of 461,986 participants investigating the association between meat intake and common cancer sites, total, red and processed meat intakes were positively associated with risks of colorectal cancer. Additionally, red meat intake was positively associated with breast cancer and prostate cancer, while poultry intake was positively associated with the risk for cancers of the lymphatic and hematopoietic tissues. However, when accounting for multiple testing using Bonferroni correction, only the associations between intake of total, red and processed meat and colorectal cancer risk, and between intake of red and processed meat and colon cancer risk were robust [21].

We found that a 70 g/day higher red and processed meat intake was associated with a 31% greater risk of colorectal cancer and 38% greater risk of colon cancer. This association was in the same direction but of greater magnitude than the results from the WCRF/AICR meta-analysis (Risk ratio (RR) for colorectal cancer 1.12, 95%-CI 1.04-1.21 per 100g/day higher red and processed meat intake), and most more recent research, including a previous study in UK Biobank investigating associations between diet and colorectal cancer with a shorter follow-up and fewer cases [19, 22-27]. Comparing the associations with colorectal cancer risk by each meat type, suggests that the association between total meat intake and colorectal cancer was driven by red and processed meat and not by poultry intake. In this study, associations with red meat intake were robust to restricting the sample to never smokers, but associations between processed meat intake and colon cancer disappeared. Most earlier studies that investigated differences in the association between red and or processed intake and colorectal cancer by smoking status reported no differences [28-30], and one study in a Japanese population showed a stronger association between processed meat intake and colon cancer in non-smokers [31]. Furthermore, in the current study associations were only significant in men although there was a suggestive similar direction for risk of colon cancer in women; there was an inverse association between poultry intake and rectal cancer risk in women and a positive association in men. The heterogeneity by sex found in our study was weaker than reported earlier in the same cohort with a smaller number of cases [19]. Women in UK Biobank were less likely to consume meat frequently and had a lower incidence of colorectal cancer, which might have made it more difficult to detect an association. Nevertheless, further investigation of the sex difference is warranted since the difference in risk, especially for rectal cancer, was substantial.

In contrast to the findings from the WCRF/AICR, in our study associations with colon cancer were slightly stronger for red meat intake than for processed meat intake. This might be explained by differences in the definition and composition of processed meat between published studies which could affect the range of carcinogenic compounds present. In our study, processed meat included processed white meat, which was not the case in some earlier studies that included red processed meat only; where white meat was included, the proportion of processed meat which was white processed meat is likely to differ by population [22, 32, 33]. Furthermore, red meat eaters and processed meat eaters overlapped strongly, with those reporting high intakes of red meat also reporting high intakes of processed meat. Therefore it might not be possible to differentiate the relative strength of risk associations between red and processed meat. In our study processed meat was more strongly associated with rectal cancer than with colon cancer. This was in contrast with most earlier studies [22], except for one recent study in a Dutch cohort that found an association between processed meat intake and rectal cancer that was similarly strong to what was found in the present study (HR per 25 g/day higher intake 1.36, 95%-CI 1.01-1.81) [27]; this might be explained by reasons similar to those described above.

Several pathways have been suggested by which both red meat and processed meat could increase the risk of colorectal cancer. Haem iron in red meat can catalyse the N-nitrosation of amines and amides to N-Nitroso compounds (NOCs) endogenously, which can generate mutations in the gastro-intestinal mucosa [34-37]. In experimental studies, haem iron has furthermore been shown to lead to lipid peroxidation that could induce genetic mutations in intestinal epithelial cells, and to affect the gut microbiota in ways which could trigger tumorigenesis through low-grade inflammation [36, 38]. Meat processing and cooking can additionally result in the formation of polycyclic aromatic hydrocarbons (PAHs) and heterocyclic amines (HCAs), which may increase the risk of colorectal cancer [39, 40]. Furthermore, processed red meat can contain NOCs from the addition of sodium nitrite and nitrates for preservation [40-42].

Although we found positive associations of total, red and processed meat intake with lung cancer, these results may be due to residual confounding by smoking, as these associations were null in never smokers. Very few cohort studies have investigated associations between meat intake and lung cancer stratified by smoking status, and results from case-control studies that suggest an apparent association in never smokers could be affected by recall bias [43]. Given that smoking is a very strong risk factor for lung cancer, it is likely that statistical adjustment for smoking status is insufficient to control for confounding by smoking, which is positively correlated with meat intake. Future studies reporting findings stratified by smoking status are needed to clarify whether any true association between meat and lung cancer exists or whether the apparent associations are entirely due to residual confounding.

We also found that red meat intake was positively associated with breast cancer and prostate cancer risk, and poultry intake with the risk for cancers of the lymphatic and hematopoietic tissues; however, these associations were not robust against correction for multiple testing. The associations with breast cancer were broadly similar to that found in the WCRF meta-analysis [44]. Of three more recent studies that also found positive associations [17, 45, 46], two considered these associations non-significant as a result of stricter significance criteria [17, 46]. It should be noted that our findings contrast somewhat with earlier research by Anderson et al. in the same cohort (UK Biobank), that found no association between red meat intake and breast cancer risk, but did find an association with processed meat intake [47]. We believe the differences from the current study are most likely due to a longer follow-up and larger number of cases in our study, but may also be due to differences in dietary data processing and analyses. The association in the current study of red meat with prostate cancer risk is consistent with some but not all previous research; the WCRF/AICR meta-analysis and more recent studies found no association between red meat intake and prostate cancer risk, while the IARC review noted a possible positive association with prostate cancer risk [5, 45, 48]. Finally, the association between poultry intake and cancers of the lymphatic and hematopoietic tissues, which seemed to be driven mainly by NHL, was consistent with results from the European Prospective Investigation into Cancer and Nutrition in which higher poultry intake was associated with an increased risk of NHL [49]. Yet, two US studies found no association between poultry intake and NHL [50, 51]. Further research in large cohorts is needed to investigate the association between meat intake and these cancer sites.

### Strengths and limitations

Major strengths of this study are the prospective design and the large sample size, which allowed investigation of a wide range of cancer sites chosen *a priori* to eliminate outcome selection bias. Data linkage to cancer registries reduced the risk of outcome misclassification and selective drop-out. Random error in dietary intakes was reduced by calibrating intakes using repeat measurements from 24h recall data to estimate usual dietary intakes in each intake category [16, 17]. Furthermore, associations were robust after adjustment for a wide range of potential confounders. Finally, the risk of reverse causation was reduced in sensitivity analyses excluding the first 2 years of follow-up, and the risk of residual confounding by smoking was addressed in sensitivity analyses restricted to never smokers.

Nevertheless, limitations need to be considered in the interpretation of our results. This study was not representative of the UK population, therefore selection bias might still have some impact on the findings, but exposure-disease relationships are likely to be generalizable because they have been shown to not require representativeness [13, 52]. Although the study was based on large case numbers for the most common cancers, it is possible that we did not have enough power to detect associations in less common cancer sites. Also, this study did not consider subtypes of cancer sites such as subsites in stomach cancer or tumour characteristics, as this information is not yet available in UK Biobank. Another limitation was the method of dietary assessment; the touchscreen questionnaire did not allow the calculation of total energy intake. We accounted for confounding by other dietary factors by adjusting for total fruit, vegetable and cereal fibre intake, and models were adjusted for BMI and physical activity [53]; however residual confounding could still operate [54].

### Conclusions

The present study supports earlier findings of an association between red and processed meat intake and colorectal cancer. More research is needed regarding the positive associations between poultry intake and cancers of lymphatic and hematopoietic tissues, and between red meat intake and risks of breast and prostate cancer.

## Data Availability

This research has been conducted using the UK Biobank Resource under application number 24494. All bona fide researchers can apply to use the UK Biobank resource for health related research that is in the public interest.

## Abbreviations

AICR: American Institute for Cancer Research
BMI: Body mass index
CI: Confidence interval
HR: Hazard ratio
HRT: Hormone-replacement therapy
IARC: International Agency for Research on Cancer
ICD: International Statistical Classification of Diseases
MET: Metabolic equivalents.
OCP: Oral contraceptive pill
RR: Risk ratio
SD: Standard deviation
WCRF: World Cancer Research Fund

## Acknowledgements

This research has been conducted using the UK Biobank Resource under application number 24494. We thank all participants, researchers and support staff who make the study possible.

## References

1. Insitute for Health Metrics and Evaluation (IHME). GBD Compare Seattle, WA: IHME, University of Washington; 2017 [01/03/2019]. Available from: http://ihmeuw.org/4qdk.

2. Ferlay J, Ervik M, Lam F, Colombet M, Mery L, Piñeros M, et al. Global Cancer Observatory: Cancer Today. Lyon, France: International Agency for Research on Cancer. 2018 [28/02/2019]. Available from: https://gco.iarc.fr/today.

3. Department of Health & Social Care. Policy Paper: 2010 to 2015 government policy: cancer research and treatment: GOV.UK; 2015 [01/03/2019]. Available from: https://www.gov.uk/government/publications/2010-to-2015-government-policy-cancer-research-and-treatment/2010-to-2015-government-policy-cancer-research-and-treatment.

4. WCRF/ AICR. Continuous Update Project Expert Report 2018. Meat, fish and dairy products and the risk of cancer. World Cancer Research Fund/American Institute for Cancer Research, 2018.

5. Bouvard V, Loomis D, Guyton KZ, Grosse Y, Ghissassi FE, Benbrahim-Tallaa L, et al. Carcinogenicity of consumption of red and processed meat. Lancet Oncol. 2015;16(16):1599-600. https://doi.org/10.1016/S1470-2045(15)00444-1. PMID: 26514947.

6. Lippi G, Mattiuzzi C, Cervellin G. Meat consumption and cancer risk: a critical review of published meta-analyses. Crit Rev Oncol Hematol. 2016;97:1-14. https://doi.org/10.1016/j.critrevonc.2015.11.008. PMID: 26633248.

7. Cross AJ, Leitzmann MF, Gail MH, Hollenbeck AR, Schatzkin A, Sinha R. A prospective study of red and processed meat intake in relation to cancer risk. PLOS Med. 2007;4(12):e325. https://doi.org/10.1371/journal.pmed.0040325. PMID: 18076279.

8. Daniel CR, Cross AJ, Graubard BI, Hollenbeck AR, Park Y, Sinha R. Prospective investigation of poultry and fish intake in relation to cancer risk. Cancer Prev Res. 2011;4(11):1903-11. https://dx.doi.org/10.1158%-2F1940-6207.CAPR-11-0241. PMID: 21803982.

9. Schoenfeld JD, Ioannidis JP. Is everything we eat associated with cancer? A systematic cookbook review. Am J Clin Nutr. 2012;97(1):127-34. https://doi.org/10.3945/ajcn.112.047142. PMID: 23193004.

10. VanderWeele TJ. Outcome-wide epidemiology. Epidemiology. 2017;28(3):399-402. Epub 04/04. https://doi.org/10.1097/EDE.0000000000000641. PMID: 28166102.

11. Palmer LJ. UK Biobank: bank on it. Lancet. 2007;369(9578):1980-2. https://doi.org/10.1016/S0140-6736(07)60924-6. PMID: 17574079.

12. Collins R. What makes UK Biobank special? Lancet. 2012;379(9822):1173-4. https://doi.org/10.1016/S0140-6736(12)60404-8. PMID: 22463865.

13. Fry A, Littlejohns TJ, Sudlow C, Doherty N, Adamska L, Sprosen T, et al. Comparison of sociodemographic and health-related characteristics of UK Biobank participants with those of the general population. Am J Epidemiol. 2017;186(9):1026-34. https://doi.org/10.1093/aje/kwx246. PMID: 28641372.

14. Galante J, Adamska L, Young A, Young H, Littlejohns TJ, Gallacher J, et al. The acceptability of repeat Internet-based hybrid diet assessment of previous 24-h dietary intake: administration of the Oxford WebQ in UK Biobank. Br J Nutr. 2015;115(4):681-6. Epub 12/11. https://doi.org/10.1017/S0007114515004821. PMID: 26652593.

15. Greenwood DC, Hardie LJ, Frost GS, Alwan NA, Bradbury KE, Carter M, et al. Validation of the Oxford WebQ online 24-hour dietary questionnaire using biomarkers. Am J Epidemiol. 2019. https://doi.org/10.1093/aje/kwz165. PMID: 31318012.

16. Bradbury KE, Young HJ, Guo W, Key TJ. Dietary assessment in UK Biobank: an evaluation of the performance of the touchscreen dietary questionnaire. J Nutr Sci. 2018;7:e6. Epub 02/01. https://doi.org/10.1017/jns.2017.66. PMID: 29430297.

17. Key TJ, Balkwill A, Bradbury KE, Reeves GK, Kuan AS, Simpson RF, et al. Foods, macronutrients and breast cancer risk in postmenopausal women: a large UK cohort. Int J Epidemiol. 2018:dyy238-dyy. https://doi.org/10.1093/ije/dyy238. PMID: 30412247.

18. UK Biobank. Cancer data: linkage from national cancer registries Version 1.4. 2013.

19. Bradbury KE, Murphy N, Key TJ. Diet and colorectal cancer in UK Biobank: a prospective study. Int J Epidemiol. 2019. https://doi.org/10.1093/ije/dyz064. PMID: 30993317.

20. Stata Corp. Stata Statistical Software: Release 15. College Station, TX: StataCorp LLC; 2017.

21. Bland JM, Altman DG. Multiple significance tests: the Bonferroni method. BMJ. 1995;310(6973):170. https://doi.org/10.1136/bmj.310.6973.170. PMID: 7833759.

22. WCRF/ AICR. The Associations between food, nutrition and physical activity and the risk of colorectal cancer. World Cancer Research Fund/ American Institute for Cancer Research, 2017.

23. Jones RR, DellaValle CT, Weyer PJ, Robien K, Cantor KP, Krasner S, et al. Ingested nitrate, disinfection by-products, and risk of colon and rectal cancers in the Iowa Women’s Health Study cohort. Environ Int. 2019;126:242-51. https://doi.org/10.1016/j.envint.2019.02.010. PMID: 30822653

24. Wada K, Oba S, Tsuji M, Tamura T, Konishi K, Goto Y, et al. Meat consumption and colorectal cancer risk in Japan: the Takayama study. Cancer Sci. 2017;108(5):1065-70. Epub 2017/03/04. https://doi.org/10.1111/cas.13217. PMID: 28256076.

25. Vulcan A, Manjer J, Ericson U, Ohlsson B. Intake of different types of red meat, poultry, and fish and incident colorectal cancer in women and men: results from the Malmö Diet and Cancer Study. Food Nutr Res. 2017;61(1):1341810-. https://doi.org/10.1080/16546628.2017.1341810. PMID: 28804436.

26. Bernstein AM, Song M, Zhang X, Pan A, Wang M, Fuchs CS, et al. Processed and unprocessed red meat and risk of colorectal cancer: analysis by tumor location and modification by time. PLoS One. 2015;10(8):e0135959. Epub 2015/08/26. https://doi.org/10.1371/journal.pone.0135959. PMID: 26305323.

27. Gilsing AMJ, Schouten LJ, Goldbohm RA, Dagnelie PC, van den Brandt PA, Weijenberg MP. Vegetarianism, low meat consumption and the risk of colorectal cancer in a population based cohort study. Sci Rep. 2015;5:13484-. https://doi.org/10.1038/srep13484. PMID: 26316135.

28. Flood A, Velie EM, Sinha R, Chaterjee N, Lacey JV, Jr., Schairer C, et al. Meat, fat, and their subtypes as risk factors for colorectal cancer in a prospective cohort of women. Am J Epidemiol. 2003;158(1):59-68. Epub 2003/07/02. https://doi.org/10.1093/aje/kwg099. PMID: 12835287.

29. Chao A, Thun MJ, Connell CJ, McCullough ML, Jacobs EJ, Flanders WD, et al. Meat consumption and risk of colorectal cancer. JAMA. 2005;293(2):172-82. Epub 2005/01/13. https://doi.org/10.1001/jama.293.2.172. PMID: 15644544.

30. Singh PN, Fraser GE. Dietary risk factors for colon cancer in a low-risk population. Am J Epidemiol. 1998;148(8):761-74. Epub 1998/10/24. https://doi.org/10.1093/oxfordjournals.aje.a009697. PMID: 9786231.

31. Takachi R, Tsubono Y, Baba K, Inoue M, Sasazuki S, Iwasaki M, et al. Red meat intake may increase the risk of colon cancer in Japanese, a population with relatively low red meat consumption. Asia Pac J Clin Nutr. 2011;20(4):603-12. Epub 2011/11/19. PMID: 22094846.

32. Norat T, Bingham S, Ferrari P, Slimani N, Jenab M, Mazuir M, et al. Meat, fish, and colorectal cancer risk: the European Prospective Investigation into cancer and nutrition. J Natl Cancer Inst. 2005;97(12):906-16. https://doi.org/10.1093/jnci/dji164. PMID: 15956652.

33. English DR, MacInnis RJ, Hodge AM, Hopper JL, Haydon AM, Giles GG. Red meat, chicken, and fish consumption and risk of colorectal cancer. Cancer Epidemiol Biomarkers Prev. 2004;13(9):1509-14. PMID: 15342453.

34. Bingham SA, Hughes R, Cross AJ. Effect of white versus red meat on endogenous N-nitrosation in the human colon and further evidence of a dose response. J Nutr. 2002;132(11 Suppl):3522s-5s. Epub 2002/11/08. https://doi.org/10.1093/jn/132.11.3522S. PMID: 12421881.

35. Cross AJ, Pollock JR, Bingham SA. Haem, not protein or inorganic iron, is responsible for endogenous intestinal N-nitrosation arising from red meat. Cancer Res. 2003;63(10):2358-60. Epub 2003/05/17. PMID: 12750250.

36. Gamage SMK, Dissabandara L, Lam AK-Y, Gopalan V. The role of heme iron molecules derived from red and processed meat in the pathogenesis of colorectal carcinoma. Crit Rev Oncol Hematol. 2018;126:121-8. https://doi.org/10.1016/j.critrevonc.2018.03.025. PMID: 29759553.

37. Bastide NM, Pierre FHF, Corpet DE. Heme iron from meat and risk of colorectal cancer: a meta-analysis and a review of the mechanisms involved. Cancer Prev Res. 2011;4(2):177. https://doi.org/10.1158/1940-6207.CAPR-10-0113. PMID: 21209396.

38. Terzić J, Grivennikov S, Karin E, Karin M. Inflammation and colon cancer. Gastroenterology. 2010;138(6):2101-14.e5. https://doi.org/10.1053/j.gastro.2010.01.058. PMID: 20420949.

39. Chiavarini M, Bertarelli G, Minelli L, Fabiani R. Dietary intake of meat cooking-related mutagens (HCAs) and risk of colorectal adenoma and cancer: a systematic review and meta-analysis. Nutrients. 2017;9(5). Epub 2017/05/20. https://doi.org/10.3390/nu9050514. PMID: 28524104.

40. Santarelli RL, Pierre F, Corpet DE. Processed meat and colorectal cancer: a review of epidemiologic and experimental evidence. Nutr Cancer. 2008;60(2):131-44. https://doi.org/10.1080/01635580701684872. PMID: 18444144.

41. Lijinsky W. N-Nitroso compounds in the diet. Mutat Res Genet Toxicol Environ Mutagen. 1999;443(1):129-38. https://doi.org/10.1016/S1383-5742(99)00015-0. PMID: 10415436.

42. Bingham SA, Pignatelli B, Pollock JRA, Ellul A, Malaveille C, Gross G, et al. Does increased endogenous formation of N-nitroso compounds in the human colon explain the association between red meat and colon cancer? Carcinogenesis. 1996;17(3):515-23. https://doi.org/10.1093/carcin/17.3.515. PMID: 8631138.

43. Gnagnarella P, Caini S, Maisonneuve P, Gandini S. Carcinogenicity of high consumption of meat and lung cancer risk among non-smokers: a comprehensive meta-analysis. Nutr Cancer. 2018;70(1):1-13. Epub 2017/10/11. https://doi.org/10.1080/01635581.2017.1374420. PMID: 29016198.

44. WCRF/ AICR. The Associations between food, nutrition and physical activity and the risk of breast cancer. World Cancer Research Fund/ American Institute for Cancer Research, 2017.

45. Diallo A, Deschasaux M, Latino-Martel P, Hercberg S, Galan P, Fassier P, et al. Red and processed meat intake and cancer risk: results from the prospective NutriNet-Santé cohort study. Int J Cancer. 2018;142(2):230-7. https://doi.org/10.1002/ijc.31046. PMID: 28913916.

46. Dunneram Y, Greenwood DC, Cade JE. Diet and risk of breast, endometrial and ovarian cancer: UK Women’s Cohort Study. Br J Nutr. 2018:1-11. Epub 12/11. https://doi.org/10.1017/S0007114518003665. PMID: 30526696.

47. Anderson JJ, Darwis NDM, Mackay DF, Celis-Morales CA, Lyall DM, Sattar N, et al. Red and processed meat consumption and breast cancer: UK Biobank cohort study and meta-analysis. Eur J Cancer. 2018;90:73-82. https://doi.org/10.1016/j.ejca.2017.11.022. PMID: 29274927.

48. Wu K, Spiegelman D, Hou T, Albanes D, Allen NE, Berndt SI, et al. Associations between unprocessed red and processed meat, poultry, seafood and egg intake and the risk of prostate cancer: a pooled analysis of 15 prospective cohort studies. Int J Cancer. 2016;138(10):2368-82. https://doi.org/10.1002/ijc.29973. PMID: 26685908.

49. Rohrmann S, Linseisen J, Jakobsen MU, Overvad K, Raaschou-Nielsen O, Tjonneland A, et al. Consumption of meat and dairy and lymphoma risk in the European Prospective Investigation into Cancer and Nutrition. Int J Cancer. 2011;128(3):623-34. https://doi.org/10.1002/ijc.25387. PMID: 20473877.

50. Daniel CR, Sinha R, Park Y, Graubard BI, Hollenbeck AR, Morton LM, et al. Meat intake is not associated with risk of non-Hodgkin lymphoma in a large prospective cohort of U.S. men and women. J Nutr. 2012;142(6):1074-80. Epub 2012/04/27. https://doi.org/10.3945/jn.112.158113. PMID: 22535761.

51. Chiu BC, Cerhan JR, Folsom AR, Sellers TA, Kushi LH, Wallace RB, et al. Diet and risk of non-Hodgkin lymphoma in older women. JAMA. 1996;275(17):1315-21. Epub 1996/05/01. PMID: 8614116.

52. Batty GD, Shipley M, Tabak A, Singh-Manoux A, Brunner E, Britton A, et al. Generalizability of occupational cohort study findings. Epidemiology. 2014;25(6):932-3. Epub 2014/09/30. https://doi.org/10.1097/EDE.0000000000000184. PMID: 25265141.

53. Jakes RW, Day NE, Luben R, Welch A, Bingham S, Mitchell J, et al. Adjusting for energy intake--what measure to use in nutritional epidemiological studies? Int J Epidemiol. 2004;33(6):1382-6. https://doi.org/10.1093/ije/dyh181. PMID: 15333618.

54. Fewell Z, Davey Smith G, Sterne JAC. The impact of residual and unmeasured confounding in epidemiologic studies: a simulation study. Am J Epidemiol. 2007;166(6):646-55. https://doi.org/10.1093/aje/kwm165. PMID: 17615092.

